# SARS-CoV-2 variants associated with vaccine breakthrough in the Delaware Valley through summer 2021

**DOI:** 10.1101/2021.10.18.21264623

**Authors:** Andrew D. Marques, Scott Sherrill-Mix, John Everett, Shantan Reddy, Pascha Hokama, Aoife M. Roche, Young Hwang, Abigail Glascock, Samantha A. Whiteside, Jevon Graham-Wooten, Layla A. Khatib, Ayannah S. Fitzgerald, Ahmed M. Moustafa, Colleen Bianco, Swetha Rajagopal, Jenna Helton, Regan Deming, Lidiya Denu, Azad Ahmed, Eimear Kitt, Susan E. Coffin, Claire Newbern, Josh Chang Mell, Paul J. Planet, Nitika Badjatia, Bonnie Richards, Zi-Xuan Wang, Carolyn C. Cannuscio, Katherine M. Strelau, Anne Jaskowiak-Barr, Leigh Cressman, Sean Loughrey, Arupa Ganguly, Michael D. Feldman, Ronald G. Collman, Kyle G. Rodino, Brendan J. Kelly, Frederic D. Bushman

**Affiliations:** Department of Microbiology, Perelman School of Medicine, University of Pennsylvania, Philadelphia, PA; Pulmonary, Allergy and Critical Care Division; Department of Medicine; University of Pennsylvania Perelman School of Medicine; Philadelphia, PA; Division of Infectious Diseases, Children’s Hospital of Philadelphia, Philadelphia, PA; Division of Gastroenterology, Hepatology & Nutrition, Children’s Hospital of Philadelphia, Philadelphia, PA; Division of COVID-19 Containment, Philadelphia Department of Public Health, Philadelphia, PA; Department of Microbiology & Immunology, Center for Genomic Sciences, Drexel University College of Medicine. Philadelphia, PA; Department of Pediatrics, Perelman School of Medicine, University of Pennsylvania, Philadelphia, PA; Sackler Institute for Comparative Genomics, American Museum of Natural History, New York, NY; Molecular & Genomic Pathology Laboratory, Thomas Jefferson University Hospital, Philadelphia, PA; Jefferson Occupational Health Network for Employees and Students (JOHN), Thomas Jefferson University, Philadelphia, PA; Department of Anatomy, Pathology, and Cell Biology, Thomas Jefferson University Hospital, Philadelphia, PA; Leonard Davis Institute of Health Economics, University of Pennsylvania, Philadelphia, PA; Department of Family Medicine and Community Health, University of Pennsylvania, Philadelphia, PA; Division of Infectious Diseases; Department of Medicine & Department of Biostatistics, Epidemiology, and Informatics; Perelman School of Medicine, University of Pennsylvania, Philadelphia, PA; Department of Genetics, Perelman School of Medicine, University of Pennsylvania, Philadelphia, PA; Department of Pathology and Laboratory Medicine, Perelman School of Medicine, University of Pennsylvania, Philadelphia, PA

**Keywords:** SARS-CoV-2, COVID-19, coronavirus, genome sequencing, Philadelphia

## Abstract

The severe acute respiratory coronavirus-2 (SARS-CoV-2) is the cause of the global outbreak of COVID-19. Evidence suggests that the virus is evolving to allow efficient spread through the human population, including vaccinated individuals. Here we report a study of viral variants from surveillance of the Delaware Valley, including the city of Philadelphia, and variants infecting vaccinated subjects. We sequenced and analyzed complete viral genomes from 2621 surveillance samples from March 2020 to September 2021 and compared them to genome sequences from 159 vaccine breakthroughs. In the early spring of 2020, all detected variants were of the B.1 and closely related lineages. A mixture of lineages followed, notably including B.1.243 followed by B.1.1.7 (alpha), with other lineages present at lower levels. Later isolations were dominated by B.1.617.2 (delta) and other delta lineages; delta was the exclusive variant present by the last time sampled. To investigate whether any variants appeared preferentially in vaccine breakthroughs, we devised a model based on Bayesian autoregressive moving average logistic multinomial regression to allow rigorous comparison. This revealed that B.1.617.2 (delta) showed three-fold enrichment in vaccine breakthrough cases (odds ratio of 3; 95% credible interval 0.89-11). Viral point substitutions could also be associated with vaccine breakthroughs, notably the N501Y substitution found in the alpha, beta and gamma variants (odds ratio 2.04; 95% credible interval of 1.25-3.18). This study thus provides a detailed picture of viral evolution in the Delaware Valley and a geographically matched analysis of vaccine breakthroughs; it also introduces a rigorous statistical approach to interrogating enrichment of viral variants.

**Importance:** SARS-CoV-2 vaccination is highly effective at reducing viral infection, hospitalization and death. However, vaccine breakthrough infections have been widely observed, raising the question of whether particular viral variants or viral mutations are associated with breakthrough. Here we report analysis of 2621 surveillance isolates from people diagnosed with COVID-19 in the Delaware Valley in South Eastern Pennsylvania, allowing rigorous comparison to 159 vaccine breakthrough case specimens. Our best estimate is a three-fold enrichment for some lineages of delta among breakthroughs, and enrichment of a notable spike substitution, N501Y. We introduce statistical methods that should be widely useful for evaluating vaccine breakthroughs and other viral phenotypes.

## Introduction

The global COVID-19 pandemic is caused by infection with the virus SARS-CoV-2 (1). Analysis of whole genome sequences from global viral samples shows ongoing changes in the composition of viral populations. RNA viruses have high mutation rates, so sequence change is expected in the viral genome due to random genetic drift (2). However, selection for efficient immune evasion and transmission between humans now seem likely to be major drivers of SARS-CoV-2 diversification (3-5).

Widespread vaccination against SARS-CoV-2 was introduced in the United States in the winter of 2020-2021. First to be implemented were vaccines based on modified mRNAs, followed by adenovirus vector delivery. Vaccines are highly protective against infection, severe disease, and death. However, infection of vaccinated individuals has been widely detected, albeit typically with much milder disease course as compared to that experienced by unvaccinated individuals (6-8). Thus interest turns to the question of which viral features are associated with vaccine breakthrough infections (7-9).

Several criteria can be applied to assessing whether sequence changes in a new variant have likely evolved to promote infection and vaccine breakthrough. Substitutions found in viral spike proteins can be tested in laboratory experiments to determine whether they promote more efficient replication in human cells or reduce binding of human antibodies (10-19). Other substitutions may alter epitopes targeted by the cellular immune system (6, 20, 21). Viral lineages with diverse combinations of these substitution have been identified and designated variants being monitored and variants of concern (VBM/VOC) by the Centers for Disease Control and Prevention (CDC). Some of the substitutions in these variants have been detected arising independently on multiple genetic backgrounds, such as the spike substitutions N501Y, E484K or the 69-70 deletion (4, 22-25), supporting a model of convergent evolution.

One indication of selection for increased transmission in humans is that several new variants have spread globally and rapidly displaced pre-existing strains. This was first documented for the D614G substitution, which spread around the world in the Spring of 2020 and displaced most strains lacking this substitution (26-29). More recently, variants first identified in the UK (B.1.1.7 or alpha) (30, 31), South Africa (B.1.351 or beta), Brazil (P.1 or gamma), California (B.1.427 and B.1.429 or epsilon), New York (B.1.526 or iota) (32), and India (B.1.617.2 or delta) (8) have been suggested to be spreading at the expense of pre-existing viral types. Against this background, intense interest focuses on whether particular variants are more efficient at infecting vaccinated individuals.

We have investigated viral genomic evolution in the Delaware Valley, which encompasses the city of Philadelphia, in a sample that includes vaccine breakthrough cases. Our initial report on the first wave of infection in this area (33) revealed that lineages in Philadelphia most closely matched sequences derived from New York City, approximately 100 miles away, which is larger than Philadelphia and had an earlier peak in infection. We also found that in some cases different viral sequence polymorphisms could be found in the same patients from different body sites or in longitudinal samples, suggestive of ongoing evolution within infected individuals (33).

In this study, samples were collected from 2621 surveillance samples from infected individuals and 159 vaccine breakthrough cases through September 2021, and the representation of different variants compared. We found that several waves of variants rose and fell in prevalence over the course of our sampling period, by the end of sampling all genomes were identified as VOC delta lineages, and the delta lineage B.1.617.2 was potentially three-fold enriched among vaccine breakthrough samples compared to surveillance samples. The amino acid substitution N501Y, found in the alpha, beta, and gamma variants, was also notably enriched in vaccine breakthrough samples. We introduce a rigorous statistical approach, Bayesian autoregressive moving average logistic multinomial regression, that should be widely useful for assessing enrichment of viral variants while controlling for the changing background of circulating strains.

## Results

### The COVID-19 epidemic in the Delaware Valley

Sampling was carried out from March 2020 to September 2021. Over the course of the study, several waves of infection are evident as increased test positivity rates (Figure 1, light grey curve) and COVID-19-attributed hospitalizations in the city of Philadelphia (Figure 1, dark grey curve). Widespread vaccination was introduced in winter 2020-2021, reaching over 70% (in adults 18 years old and older) in Philadelphia by September 2021 (Figure 1, black curve). As is described below, during this period, SARS-CoV-2 variants alpha and delta, designated as VBM/VOC, became the overwhelming majority of genomes identified by our sequence surveys (Figure 1, green and red curves).

**Figure 1.**
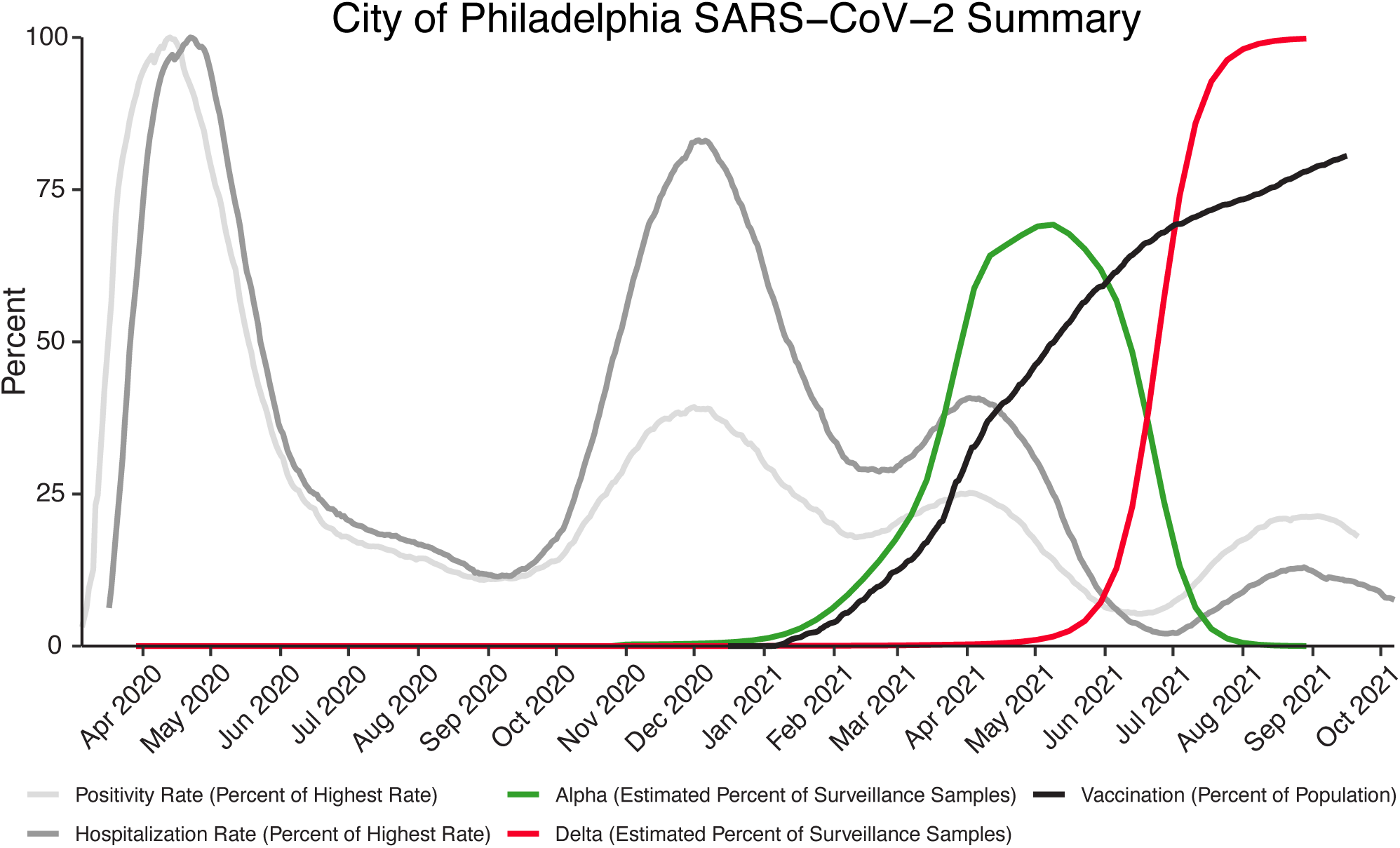
Longitudinal data from the COVID-19 pandemic in the city of Philadelphia. The y-axis shows the daily test positivity rate (light grey) as a percent of the highest value (26.57% positivity on 4/13/2020), the hospitalization rate (dark grey) as a percent of the highest value (87 hospitalizations per day on 4/22/2020), the vaccination rate in adults 18 years old and older (black). The estimated percent of surveillance samples classified as alpha (green) or delta (red) variant was estimated from the sequence data presented in this paper. Other data is from the city of Philadelphia “Testing Data: Programs and Initiatives.”

### Patient populations

Patient samples (nasal or nasopharyngeal swab and saliva) used for SARS-CoV-2 whole genome sequence analysis are listed in Table S1. To preserve patient confidentiality, samples were deidentified except for collection date and rationale for collection. Surveillance samples (n=2621) were defined as those acquired from clinical diagnostic laboratories across the Delaware Valley (n=2485), from hospitalized subjects (n=116), and asymptomatic subjects testing positive in a university screening program (n=20) (34). Vaccine breakthrough cases (n=159) were included only if detected at least 2 weeks after the final vaccine dose (second dose for Moderna and Pfizer-BioNTech mRNA vaccines or single dose for the Johnson & Johnson adenovirus vector platform) and the subject tested positive by clinical laboratory assay. Data were not available on subject immune responses following vaccination.

As a positive control, S gene target failure samples (n=172) were also compared. The TaqPath COVID-19 Combo Kit by Thermo Fisher Scientific targets three regions of the SARS-CoV-2 genome for viral detection; the ORF1ab, nucleocapsid (N), and spike (S) genes. The region of the S gene interrogated by the assay overlies a characteristic deletion in the alpha variant (del 69-70), so samples containing alpha lineage virus are selectively negative for the spike amplicon, while the other two targets are detected. Samples with these characteristics were targeted for sequencing early during the wave of alpha infections to track the variant; here these spike target gene failures serve as positive controls for the statistical model.

### Sequencing strategies

Several sequencing strategies were used to acquire whole genome sequences. The POLAR protocol with Artic primers and Illumina sequencing was used for most samples (35). Smaller numbers of samples were acquired using the Paragon, Illumina RPIP, and Illumina CovidSeq methods. Viral genome sequences were judged to be high quality and included in the study if 95% of the viral genome was covered by at least 5 reads. In all, 2952 high-quality sequences were generated and analyzed. Viral variants were assigned using Pangolin lineage software.

### Variants detected in pooled surveillance data

Figure 2A shows the proportions of variants detected in surveillance samples over the course of the study from March 2020 to September 2021. Variants detected are summarized by the color code on the bar graph (variant designations used are summarized in Table S2). The numbers of genomes analyzed per week are indicated above each column. Numbers varied both as surveillance sequencing efforts accelerated and as the availability of samples varied.

**Figure 2.**
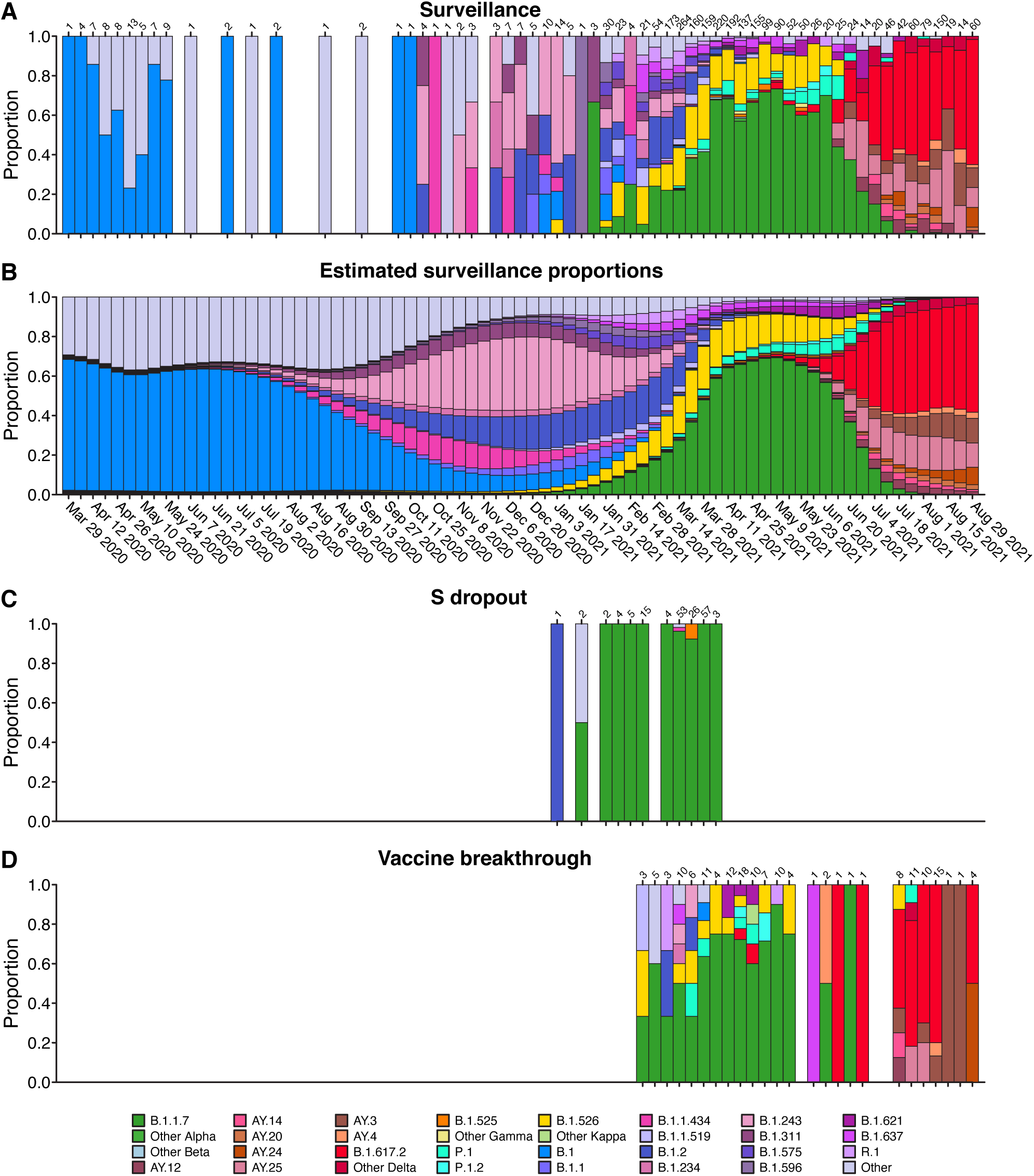
Comparison of viral genome sequence data from surveillance samples (A, B) to spike target gene failures (C) and vaccine breakthrough samples (D). A) Longitudinal stacked bar graph depicting the SARS-CoV-2 variants present in surveillance samples from the Delaware Valley, shown as the proportion of genomes classified as each variant lineage within each week. The numbers of genomes sampled each week are shown above the graph. Variants are colored according to the key at the bottom of the figure. B) Markings are the same as in A), but showing the proportions of variants estimated from the count data in A) using Bayesian autoregressive moving average multinomial logistic regression. C) Markings as in A), but showing counts of spike target gene failures samples. D) Markings as in A), but showing the counts of vaccine breakthrough samples.

Members of the B.1 lineage predominated March 2020 until fall 2020, at which time B.1.2 and B.1.243 became predominant. Later the B.1.526 and B.1.1.7 (alpha) variants emerged, with B.1.1.7 becoming predominant by spring 2021. Delta (B.1.617.2 and AY lineages) became detectable in late spring and predominant in early summer. By late summer, delta lineages were the only variants detected.

### Assessing variant abundance in the surveillance population

Wide-spread vaccination was introduced in mid-winter 2020-2021, but nevertheless breakthrough infections were detected in some vaccinated individuals, raising the question of whether specific viral features identifiable in sequence data might be associated with vaccine evasion. We sequenced 159 vaccine breakthrough samples collected between February 22 and September 3, 2021, and compared the distributions of lineages or genomic variations to those observed in surveillance samples from the same community (n=2621).

Challenges in the analysis include the facts that: 1) the distribution of variants in the surveillance samples is changing over time; 2) sampling is uneven over time; and 3) sampling is subject to stochastic fluctuations. To address these issues, we developed a model based on Bayesian analysis that combines an autoregressive moving average model with multinomial logistic regression. The underlying variant proportions at each week of the study were estimated from the counts of SARS-CoV-2 variants assuming relatively smooth changes over time and accounting for stochastic fluctuations during sampling (Figure 2B). These estimated surveillance proportions were compared to the variant counts observed in spike gene target failure samples (Figure 2C) and vaccine breakthrough samples (Figure 2D). Since the surveillance population proportions were estimated for each week, the weekly vaccine breakthrough or spike gene target failure variant counts could be compared to the corresponding time-matched surveillance estimates.

Examples of the temporal profiles of the modeled lineage succession are shown in more detail in Figure 3. One benefit of the model is that uncertainty in the surveillance estimates can be included. For example, note that time points with fewer surveillance samples (lighter grey bars) have larger 95% credible intervals (CrI) for the proportion estimate (colored shading). From this view, it is evident that several further lineages waxed and waned notably over the sampling period, including B.1.1.434 and B.1.526.

**Figure 3.**
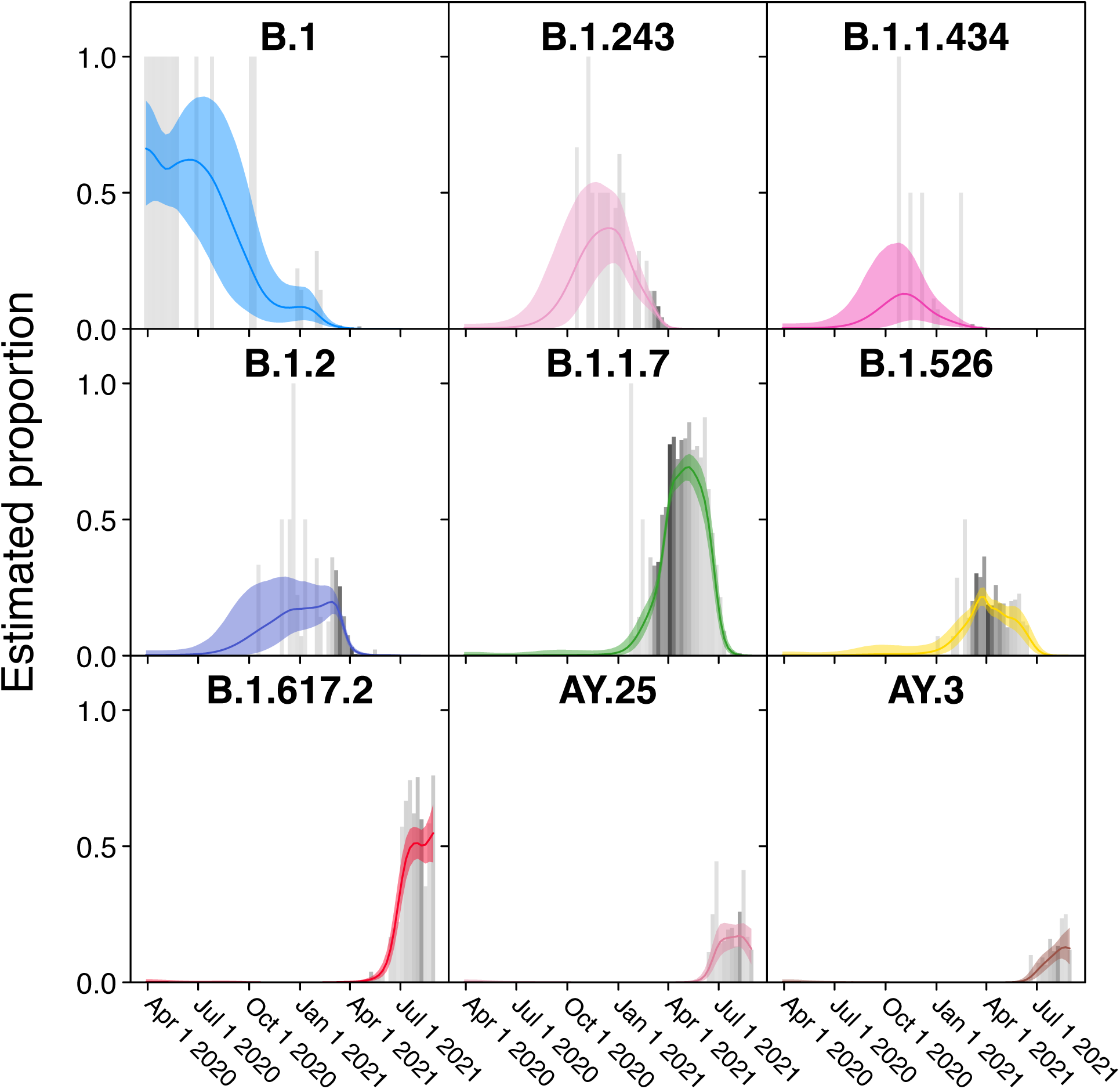
Frequencies of individual variants estimated using Bayesian autoregressive moving average multinomial logistic regression. Time is shown along the x-axis and estimated proportions of the surveillance population along the y-axis. The grey bars indicate raw proportions from the count data shaded by the number of observations observed in a given week (darker indicating more samples) while the colored lines indicate the proportion estimated by the Bayesian model. The light colored envelopes around each line show the 95% credible intervals for the proportion. Only lineages achieving an estimated proportion of >10% in any given week are shown.

### Analyzing lineages in spike gene target failures

As a control, we assessed the variants associated with spike (S) gene target failure samples (Figures 2C and 4 and Table S3), where an amplicon overlapping the 69-70 deletion in spike failed selectively. To track the spread and variation of B.1.1.7 during the alpha wave of infection, we sequenced 172 S gene target failure samples in the Delaware Valley from January to April 2021. Upon variant assignment, genomes from 96.1% of spike target gene failure samples corresponded to B.1.1.7. Rarely, other lineages were seen that share the spike deletion with B.1.1.7 (B.1.375 and B.1.525) or were likely stochastic failures of S amplification or genomes with novel combinations of mutations.

As expected, we found that the B.1.1.7 lineage was estimated to be highly enriched in the S gene target failure set over what would be expected from its frequency in the surveillance lineage counts (Figure 4) (odds ratio of 120; 95% credible interval 38-470). The B.1.525 lineage, which also contains the S69-70 deletion, was estimated to be enriched as well (odds ratio 33; 95% credible interval 1.7-380). Note that this enrichment was detected based on counts of only two B.1.525 genomes in the S gene target failure set and 12 surveillance genomes, both emphasizing the sensitivity of the model and highlighting that the detection of enrichment can be easier in lineages rare in the surveillance population. Other lineages showed little association with spike target gene failures, as expected (note that only a single B.1.375 lineage sample was observed and thus it did not reach the threshold for lineage-specific analysis and was grouped in “Other”). Thus the analysis of S target failures confirmed that the Bayesian autoregressive moving average categorical regression model was effective at identifying over-represented lineages relative to the time-adjusted expectation from surveillance.

**Figure 4.**
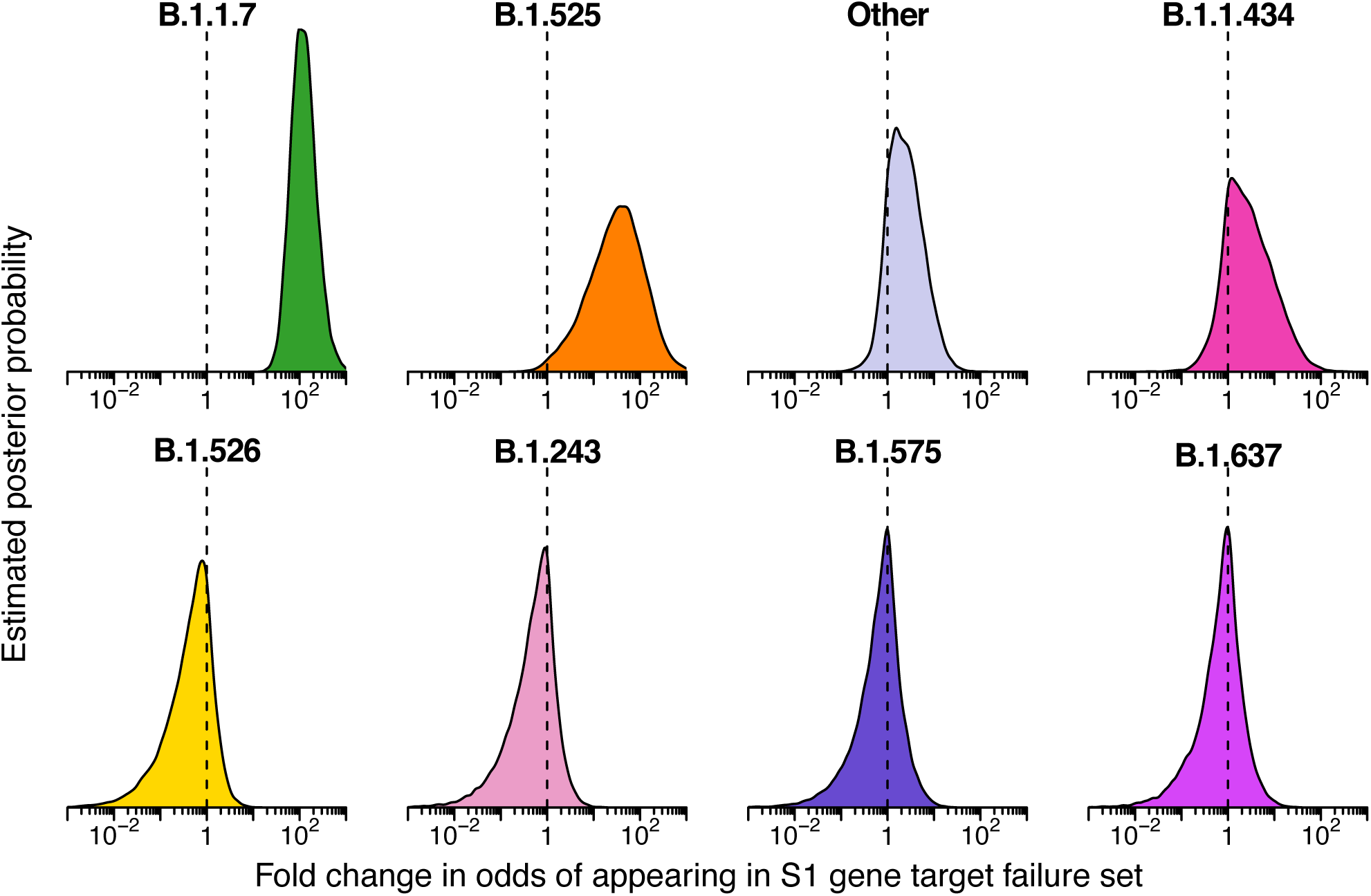
Estimated posterior probability densities for the enrichment of variants among spike gene target failures produced by Bayesian autoregressive moving average multinomial logistic regression. The x-axis shows the fold enrichment/depletion in the odds of a variant (labeled above each plot) appearing in the spike gene target failure set relative to the proportions estimated in the surveillance population and the y-axis shows the posterior probability. No enrichment (fold-change of 1) is indicated by the dashed vertical line. Increased density to the right indicates greater likelihood of inclusion among spike gene target failures, increased density to the left indicates decreased likelihood. The four lineages with highest posterior probability of enrichment are shown on the top and four lineages with high posterior probability of depletion are shown on the bottom. Color coding indicates the variant queried with colors as in earlier figures.

### Enrichment of delta variant B.1.617.2 in vaccine breakthrough samples

We then applied the Bayesian model to analyzing vaccine breakthrough samples (Figures 2D and 5 and Table S4). There was less clear enrichment in vaccine breakthroughs than in the S gene target failure set. The delta lineage B.1.617.2 did show signs of enrichment in breakthrough samples, with mean odds ratio of 3 (95% credible interval 0.89 to 11). The one-sided posterior probability of any enrichment for B.1.617.2 in vaccine breakthrough was estimated at 96% with a 73% probability of more than a 2-fold enrichment in odds of appearing in vaccine breakthrough. This enrichment was not observed for all lineages grouped within delta, with both a group-wise estimate for all delta lineages and the specific estimates for the other delta AY lineages, including AY.3, AY.4, AY.14, and AY.24, showing little enrichment over surveillance. These alternative delta lineages also had fewer samples observed, so a lack of power may be a partial explanation. Note that the power to detect enrichment of delta is limited by its rapid spread--observation of a delta vaccine breakthrough case at a time point when almost all surveillance population infections were also delta provides little information. Similarly, lineages that had already waned by the time of widespread vaccination are difficult to assess since there was little chance for them to appear in vaccine breakthrough cases.

### Assessing possible enrichment of specific mutations in spike gene target failures and vaccine breakthroughs

We next investigated whether any individual base substitutions in the viral genome were selectively associated with spike gene target failure and vaccine breakthrough samples. A summary of mutations detected is presented in Table S5. We adapted the Bayesian autoregressive moving average logistic regression method to assess the behavior of single mutations and amino acid substitutions. Figure 6 and Table S6 summarizes results. Many mutations varied in frequency over the course of the study often paralleling the profile of the most abundant lineages containing them (Figure 6A).

**Figure 5.**
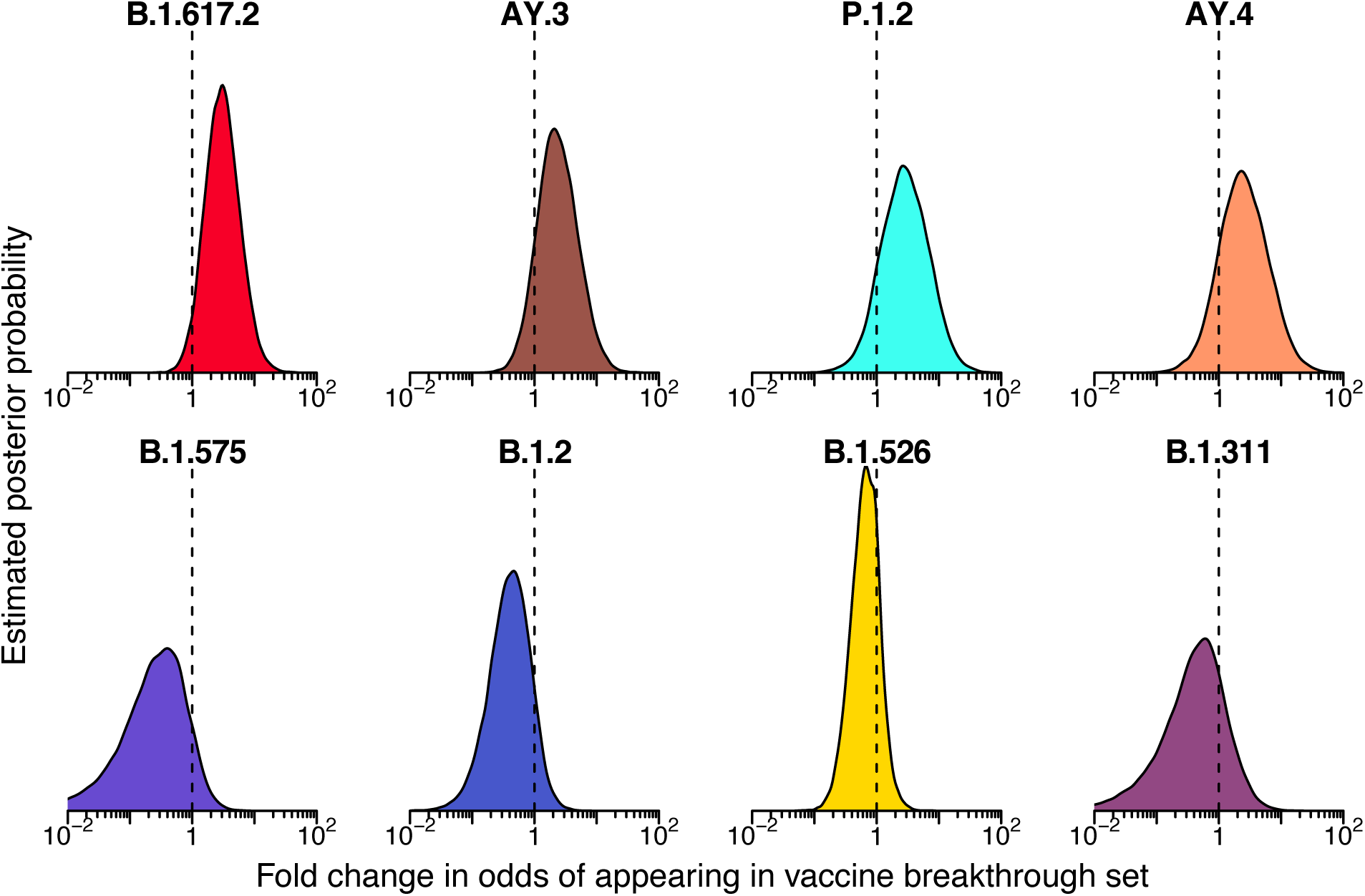
Posterior probability densities for enrichment of variants among vaccine breakthrough samples compared to the surveillance population as estimated by Bayesian autoregressive moving average multinomial logistic regression. Markings as in Figure 4.

**Figure 6.**
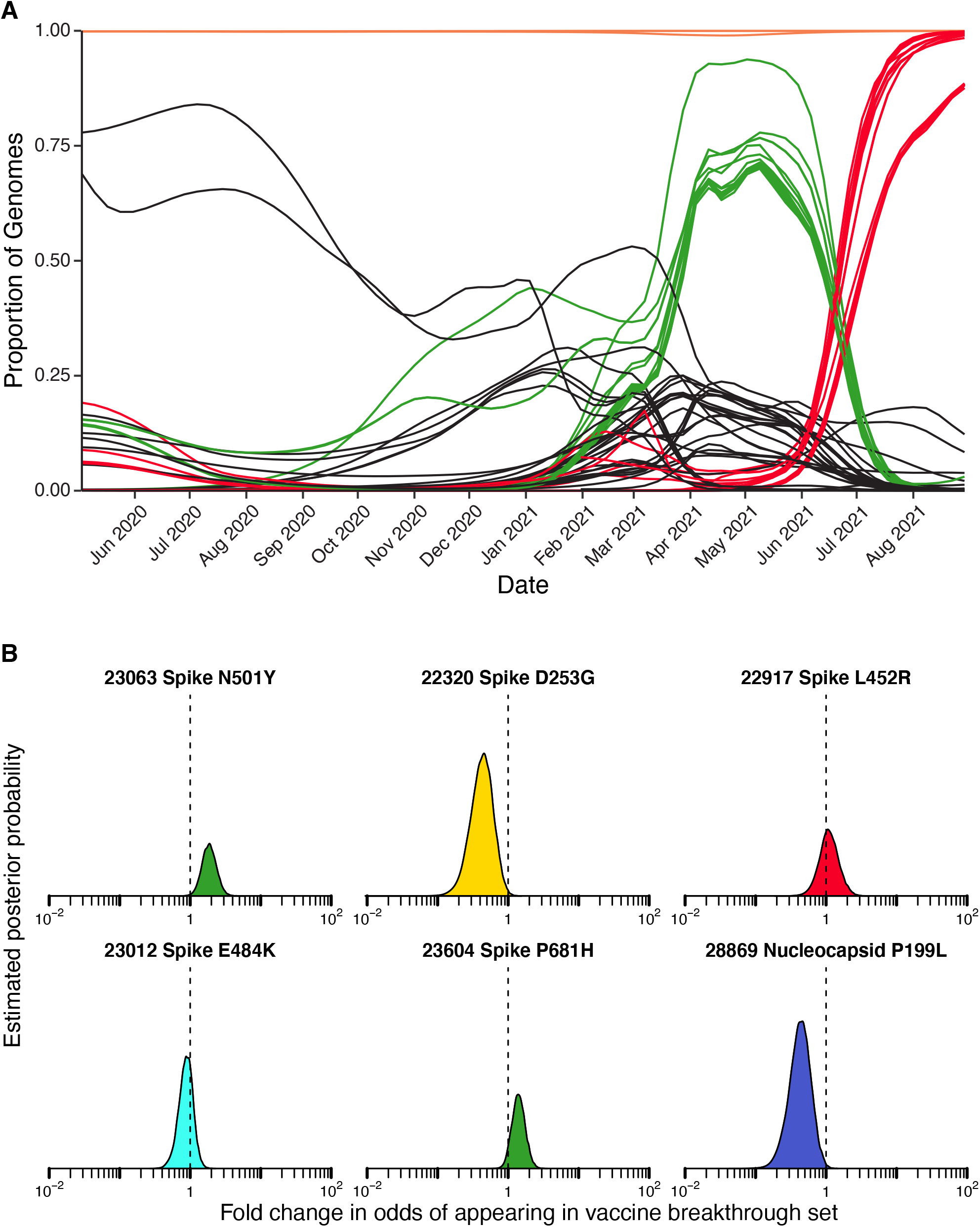
Assessment of enrichment of specific base substitutions and deletions in vaccine breakthrough samples. A) Longitudinal frequencies of individual mutations estimated using Bayesian autoregressive moving average logistic regression models. Red indicates mutations commonly found in delta lineages. Green indicates mutations commonly found in alpha lineages. Orange indicates mutations shared by almost all lineages in the study. Black indicates mutations found in other subsets of lineages. B) Estimated posterior probability densities summarizing the fold enrichment/depletion in odds of a mutation appearing in the vaccine breakthrough samples over its proportions estimated from surveillance samples. The mutations estimated as most enriched and most depleted are shown along with other mutations of interest. Markings as in Figures 4 and 5.

As expected, the 69-70 deletion was found to be highly enriched in the S gene target failure set (odds ratio of 205, 95% credible interval of 65-581). Enrichment among S gene target failures was also seen for another 28 mutations (Table S6), where all the most strongly enriched substitutions were characteristic of the alpha variant and thus due to “hitchhiker” effects. Underrepresentation in spike target gene failures was also seen in mutations in multiple additional open reading frames, reflecting mutations found in variants lacking the 69-70 deletion. Each mutation was analyzed separately without accounting for genomic linkage so this is as expected.

For vaccine breakthroughs (Figure 6B and Table S7), the most notably enriched substitution was N501Y, which showed an odds ratio of 2.04 (95% credible interval of 1.25-3.18). The N501Y substitution is found in multiple VBM, including alpha, beta, and gamma, and is reported to increase the affinity of spike protein binding to the ACE2 receptor (24, 36, 37) and to diminish binding of some human antibodies to spike (25). The spike substitution P681H was also slightly enriched; this substitution is near the furin cleavage site and may promote efficient proteolysis. Two other closely studied spike substitutions, E484K and L452R, were not notably enriched among vaccine breakthrough cases, and the D253G substitution was modestly depleted. Among non-spike substitutions, the P199L substitution in the nucleocapsid showed notable depletion (Table S7). Stepping back, these findings together emphasize the potential importance of the N501Y substitution in vaccine breakthrough.

## Discussion

Vaccination against SARS-CoV-2 has been highly effective at preventing infection, hospitalization and death. However, infections occasionally take place despite vaccination. It has been suggested that certain lineages of SARS-CoV-2 are more prone to evade vaccination, however, interpretation of counts of vaccine breakthrough is difficult due to variation over time in circulating lineages, the numbers of vaccinated individuals, times since vaccination and the application of non-pharmaceutical interventions. Here we introduce modeling based on Bayesian autoregressive moving average multinomial logistic regression, which estimates the proportions of lineages in the surveillance data over time, allowing comparison of lineage counts in populations of interest to time-matched surveillance estimates. Multiple studies have investigated whether certain lineages are more likely to appear in vaccine breakthrough infections (7, 8, 38-42). However, few studies have as large numbers of sequenced samples for both background surveillance and for vaccine breakthrough cases within a matched geographical region as reported here. Thus our contribution provides targeted data on the nature of breakthroughs at nucleotide resolution.

Using this method, we found that there was evidence for enrichment in the odds of variant lineages appearing in the vaccine breakthrough population relative to the general surveillance population, with the delta variant B.1.617.2 showing the strongest signal with an estimated enrichment of 3-fold (95% credible interval 0.89 to 11). Several previous studies have also noted delta variants to be enriched among vaccine breakthroughs (7, 8); our data provides further support based on careful statistical analysis.

Using a similar Bayesian autoregressive moving average logistic regression model, we interrogated enrichment of point substitutions among vaccine breakthrough cases. The N501Y substitution stood out as particularly associated with vaccine breakthroughs. N501Y is found in several of the VBM, and this substitution is well known to increase affinity of binding for the viral spike protein to the ACE2 receptor (24, 36, 37), as well as reduce antibody binding to promote immune evasion (25). Recently the N501Y substitution was suggested to be a central feature in a meta-signature of up to 35 mutations which recur in alpha, beta, gamma and other lineages, and which mark a viral fitness peak reflecting optimization for spread in humans (43). In contrast, several substitutions were estimated to be less likely to appear in vaccine breakthrough samples, including nucleocapsid mutation P199L (detected in B.1.2, B.1.526, and B.1.596; odds ratio of 0.5; 95% credible interval of 0.24-0.88), and spike mutation D253G (detected in B.1.526; odds ratio of 0.5; 95% credible interval of 0.23-0.89). The spike D253G substitution has been implicated in mAb evasion (44), and nucleocapsid P199L may alter the assembly of SARS-CoV-2 VLPs (45), functions possibly linked to their depletion in vaccine breakthrough.

This study has several limitations. For the vaccine breakthrough samples, we did not have paired immunological data, so it is unknown whether the vaccinations provoked protective immune responses. Our surveillance data was from a mixture of hospitalized patients, symptomatic subjects tested in clinical diagnostic laboratories, and asymptomatic subjects tested weekly at an academic institution, and so our sampling may not be entirely representative of viruses circulating in the community. Several different amplification and sequencing methods were used to acquire data. Documentation of vaccination status may be incomplete. For all the viral genome sequencing, only samples achieving a threshold level of viral RNA could be sequenced (roughly Ct <28 for swab-based testing, Ct <20 for saliva), so it is possible that other variants predominate in subjects with lower viral loads. Studies have been initiated to address some of these concerns.

In summary, similar to other regions in the US and around the world, VBM/VOC and other lineages waxed and waned in the Delaware Valley, with delta lineages ultimately comprising all recent samples in the fall of 2021. Widespread vaccination was introduced in the region in winter 2020-2021, and increasing numbers of breakthrough infections have since been detected. To compare the lineages found in breakthrough with those observed in general surveillance, we introduce analysis based on Bayesian autoregressive moving average multinomial logistic regression, and show that delta variant B.1.617.2 showed 3-fold enrichment in vaccine breakthroughs. The N501Y substitution stood out among point substitutions for enrichment in vaccine breakthroughs. We expect that these modeling methods will be useful in monitoring the effectiveness of vaccination programs going forward as novel variants of SARS-CoV-2 continue to emerge.

## Materials and Methods

### Human subjects

The University of Pennsylvania Institutional Review Board (IRB) reviewed the research protocol and deemed the limited data elements extracted with positive SARS-CoV-2 specimens to be exempt from human subject research per 45 CFR 46.104, category 4 (IRB #848605). For hospitalized subjects at the University of Pennsylvania, following informed consent (IRB protocol #823392), patients were sampled by collection of saliva, oropharyngeal and/or nasopharyngeal swabs, or endotracheal aspirates if intubated, as previously described (33). Clinical data was extracted from the electronic medical record. Further samples were collected from asymptomatic subjects detected in a screening program at the Perelman School of Medicine at the University of Pennsylvania and symptomatic subjects tested throughout the PennMedicine clinical network under IRB protocols #843565 and #848608. Human samples were collected at Children’s Hospital of Philadelphia under protocol # 21-018726 approved by the Children’s Hospital of Philadelphia IRB. Human samples were collected at Thomas Jefferson University under protocol number IRB Control # 21E.441 approved by the Thomas Jefferson University IRB. Vaccine breakthrough cases were identified by either report to HUP Infection and Control, Jefferson Infection and Control, CHOP Department of Infection Control and Prevention, the Philadelphia Department of Public Health, or, where applicable, chart review.

### Sequencing methods

Several sequencing methods were used to acquire viral whole genome sequences.

The POLAR protocol was used to acquire the majority of viral genome sequences (46). Illumina’s NextSeq instrument was used to gather sequence data. In detail, 5 μl of viral RNA, 0.5 μl of 50 μM Random Hexamers (Thermo Fisher, N8080127), 0.5μl of 10mM dNTPs Mix (Thermo Fisher, 18427013), and 1 μl nuclease-free water was incubated at for 5 minutes at 65ºC proceeded by a 1 minute incubation at 4ºC. To perform reverse transcription, 6.5 μl from the previous reaction, 0.5 μl SuperScript III Reverse Transcriptase (Thermo Fisher, 18080085), 0.5 μl of 0.1M DTT (Thermo Fisher, 18080085), 0.5 μl of RNaseOut (Thermo Fisher, 18080051), and 2 μl of 5X First-Strand Buffer (Thermo Fisher, 18080085) was incubated for 50 minutes at 42ºC, followed by an incubation for 10 minutes at 70ºC, and then held at 4ºC. To amplify the cDNA, artic-ncov2019 version 3 primers were used (IDT). To perform PCR amplification of the viral cDNA, the following reagents were added to 2.5 μl of the previous mixture: 0.25 μl Q5 Hot Start DNA Polymerase (NEB, M0493S), 5 μl of 5X Q5 Reaction Buffer (NEB, M0493S), 0.5 μl of 10mM dNTPs Mix (NEB, N0447S), either 4.0 μl of pooled primer set 1 or 3.98 μl of pooled primer set 2, and nuclease-free water to bring to a final volume of 25 μl. This PCR amplification of the viral cDNA used the following conditions: 98ºC for 30 seconds for 1 cycle, 25 cycles at 98ºC for 15 seconds and 65ºC for 5 minutes, and then held at 4ºC. Amplicons generated by the two primer sets from the same sample were pooled together then diluted to a concentration of 0.25 ng/μl. The Nextera library was prepared using the Nextera XT Library Preparation Kit (Illumina, FC-131-1096) and the IDT for Illumina DNA/RNA UD Indexes Set A and B (Illumina, 20027213, 20027214, 20027215, 20027216). The Quant-iT PicoGreen dsDNA quantitation assay kit was used to quantify the DNA of each sample (Invitrogen, P7589). The samples were pooled in equal quantities, and the pooled library was quantified using the Qubit1X dsDNA HS Assay Kit (Invitrogen, Q33230). The library was sequenced on an Illumina NextSeq.

Another sequencing method, used at CHOP, was Paragon. Paragon sequencing was carried out as in (23). Briefly, RNA was extracted from nasopharyngeal swab samples using QIAamp Viral RNA Mini (Qiagen). Whole genome sequencing was carried out by the Genomics Core Facility at Drexel University. Amplification was performed using Paragon Genomics CleanPlex SARS-CoV-2 Research and Surveillance NGS Panel 1 and 2. Libraries were quantified using the Qubit dsDNA HS (High Sensitivity) Assay Kit (Invitrogen) with the Qubit Fluorometer (Invitrogen). Library quality was assessed using Agilent High Sensitivity DNA Kit and the 2100 Bioanalyzer instrument (Agilent). Libraries were normalized to 5nM and pooled in equimolar concentrations. The resulting pool was quantified again using the Qubit dsDNA HS (High Sensitivity) Assay Kit (Invitrogen) and diluted to a final concentration of 4nM; libraries were denatured and diluted according to Illumina protocols and loaded on the MiSeq at 10pM. Paired-end and dual-indexed 2×150bp sequencing was carried out using MiSeq Reagent Kits v3 (300 cycles).

The Thomas Jefferson University site carried out sequencing using the Illumina RPIP and CovidSeq methods essentially as per the manufacturer’s instructions. A nasopharyngeal swab specimen that was tested positive by PCR for SARS-CoV-2 was used for this study. Vaccine breakthrough specimens were identified by Jefferson Occupational Health Network for Employees and Students (JOHN) and were sequenced with a designation of VBT. Randomly selected residual positive specimens from the Molecular & Genomic Pathology Laboratory at Jefferson during the same period were sequenced for the purpose of epidemiology surveillance and labeled as SURV. RNA was extracted from 200 μl of the specimen and eluted in 110 μl using the Biomérieux EasyMag Extraction System, following EasyMag’s Generic protocol. Whole genome sequencing for SARS-CoV-2 was subsequently performed at the Molecular & Genomic Pathology Laboratory at Thomas Jefferson University Hospital. With the exception of a few samples, samples were sequenced using Illumina COVIDSeq protocol (Illumina) following manufacturer’s guideline. Remaining specimens were sequenced using the Swift Normalase® Amplicon Panels (SNAP) Core Kit along with the SARS-CoV-2 Additional Genome Coverage Primer Panel following the manufacturer’s protocol. For data analysis, Illumina’s Local Run Manager’s GenerateFASTQ module was used to generate the fastq files for all specimens. The fastq files were transferred to UPenn’s secure data server for further processing to obtain QC metrics and lineage data.

Samples with VSP numbers lower than VSP00256 (Table S1) were previously described in Everett et al. (33).

### Data analysis

To process sequence data, sequence reads are trimmed to remove low quality base calls (< Q20) and aligned to the original Wuhan reference sequence (NC_045512.2) with the BWA aligner tool (v0.7.17) (47), after which alignments are filtered with the Samtools package (v1.10)(48) to remove suspect alignments. Sequencing depth is determined for each position in the viral genome and genomes are accepted for analysis when ≥ 95% of genome positions are covered with a read depth of ≥ 5 reads. Variant positions are called with the Bcftools package (v1.10.2-34)(49) requiring PHRED scores ≥ 20 and variant read frequencies ≥ 50% of the total reads. The nature of substitutions is determined by retrieving reading frames from the reference GENBANK record, translating, and determining the native and mutant residues.

Variants were assigned using the Pangolin lineage software (Pangolin version 3.1.11 with the PangoLEARN 2021-08-24 release). Note that these lineages are updated regularly and so are expected to change over time. Point mutations were assigned using a previously described bioinformatics pipeline (32).

Statistical analysis of variant enrichment and mutation enrichment in subsets of the data were separately assessed using a Bayesian model and Markov chain Monte Carlo sampling implemented in Stan (50). The model takes the vector of counts of the variants or mutations seen in the surveillance sampling for a given week, counts_*i*_, and assumes they are multinomially distributed where:

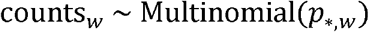

with probabilities modeled as multinomial logistic:

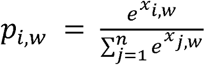

where *p*_*i,w*_ is the true proportion of variant or mutation *i* on week *w. p*_*,*w*_ indicates the vector of all lineage probabilities for week *w* and *n* is the total number of variants or mutations observed. The underlying proportions for a given week are assumed to be centered around the proportions observed in the prior week (autoregressive) plus a change term that is itself correlated with the change observed in the prior week (moving average):

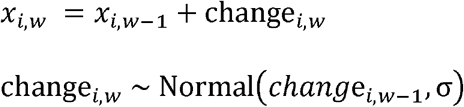

where change_*i,w*_ is the change in log odds of variant or mutation *i* on week *w*. The initial starting proportions are given flat priors:

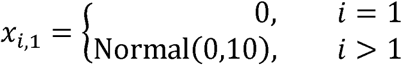

The standard deviation for changes, *σ*, was given a *Gamma*(1,2) prior distribution.

To assess if there’s enrichment of a particular variant or mutation in the vaccine breakthrough, counts of vaccine breakthrough samples for a given week, vaccineCounts_*w*_, were modeled as:

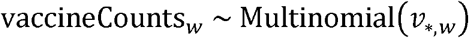

where:

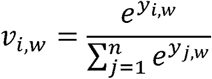

and

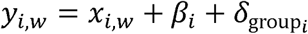

where *β*_*i*_ measures the log odds enrichment/depletion of variant or mutation *i* in the vaccine breakthrough population over the surveillance population, group_*i*_ indicates the WHO classification for Pango lineage *i* and 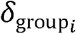 indicates the enrichment or depletion for a CDC VBM/VOC classification containing more than one Pango lineage, e.g. Delta containing B.1.617.2 and other variants with an AY prefix. In this data, Delta, Gamma and Non-VBM/VOC groupings contained more than one lineage above the abundance threshold. For WHO classifications containing only a single lineage, 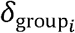 was set to 0. The β and remaining δ were given DoubleExponential(0,1) priors.

This was repeated equivalently in the samples collected as S gene target failures, with the counts, SFailCounts_*w*_, modeled as:

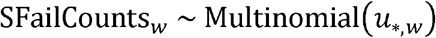

where:

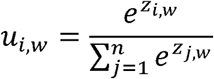

and

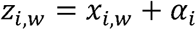

The were given DoubleExponential(0,1) priors.

For the assessment of lineage enrichment, genomes from lineages that had more than 10 genomes assigned to them were included as their individual lineages, genomes from lineages with 10 or fewer assignments that were listed within VBM/VOC were included as miscellaneous classification for their particular WHO category e.g. “Other delta” and all other genomes were grouped into an Other category. For the assessment of mutations, each mutation that was found in greater than 5% of genomes was independently modeled as above but replacing the Multinomial distributions with Binomial and removing the inapplicable lineage group *δ* terms.

### Data availability

All viral genome sequences acquired in this study have been deposited in GISAID and at NCBI under accession numbers listed in Table S1. Analysis code is available at http://github.com/sherrillmix/longitudinalLineages and will be deposited on Zenodo prior to final submission. Sequence processing software and intermediate files used in this study are available at DOI:10.5281/zenodo.5559699. A list of key reagents used in this study is in Table S8.

## Acknowledgements

We are grateful to individuals and their families who volunteered to provide specimens, members of the Bushman and Collman laboratories for help and suggestions, and to Laurie Zimmerman for artwork and help with manuscript preparation. We acknowledge help from all the staff of the Philadelphia Department of Public Health. Funding was provided by a contract award from the Centers for Disease Control and Prevention (CDC BAA 200-2021-10986 and 75D30121C11102/000HCVL1-2021-55232), philanthropic donations to the Penn Center for Research on Coronaviruses and Other Emerging Pathogens, and in part by NIH grant R61/33-HL137063 and AI140442 -supplement for SARS-CoV-2 and NIAID contract 75N93021C00015. BJK is supported by NIH K23 AI 121485. Additional assistance was provided by the Penn Center for AIDS Research (P30-AI045008) and the Women’s Committee of CHOP.

## Author contributions

ADM, JE, SS-M, RGC, KR, BJK and FDB designed the study; SAW, JG-W, LAK, ASF, ADM, CB, SR, JH, RD, LD, AA, EK, SC, CN, JCM, PP, CC, KS, AG, MF, RGC, KR, AJ-B, LC, SL, AG and BJK managed acquisition of clinical specimens; ADM, CB, SR, LD, AA, JCM, PP, NB, Z-XW, PH, AMR, AG, AM, and FDB carried out sequencing; ADM, AMM, CB, SR, AA, JCM, PP, SS-M, BJK, JE and FDB carried out bioinformatic and statistical analysis; ADM, SS-M, JE, KR, BJK, RGC, and FDB wrote the paper.

## Supplementary Material

Table S1. Human subjects and SARS-CoV-2 genome sequences analyzed in this work, including genome quality metrics, viral variant designation, and accession numbers.

Table S2. Nomenclature of VBM/VOC used in this study.

Table S3. Estimated fold enrichment in odds of appearing in the spike gene target failure set for each SARS-CoV-2 lineage.

Table S4. Estimated fold enrichment in odds of appearing in vaccine breakthrough samples for each SARS-CoV-2 lineage.

Table S5. The percent of genomes within each SARS-CoV-2 lineage containing each substitution or deletion.

Table S6. Estimated fold enrichment in odds of appearing in the spike gene target failure set for each substitution or deletion studied.

Table S7. Estimated fold enrichment in odds of appearing in the vaccine breakthrough samples for each substitution or deletion studied.

Table S8. Key reagents

## References

1. Lu R, Zhao X, Li J, Niu P, Yang B, Wu H, et al. Genomic characterisation and epidemiology of 2019 novel coronavirus: implications for virus origins and receptor binding. Lancet. 2020;395(10224):565–74.

2. Holmes EC. Error thresholds and the constraints to RNA virus evolution. Trends Microbiol. 2003;11(12):543–6.

3. Holmes EC, Goldstein SA, Rasmussen AL, Robertson DL, Crits-Christoph A, Wertheim JO, et al. The origins of SARS-CoV-2: A critical review. Cell. 2021;184(19):4848–56.

4. Meng B, Kemp SA, Papa G, Datir R, Ferreira I, Marelli S, et al. Recurrent emergence of SARS-CoV-2 spike deletion H69/V70 and its role in the Alpha variant B.1.1.7. Cell Rep. 2021;35(13):109292.

5. Kistler KE, Huddleston J, Bedford T. Rapid and parallel adaptive mutations in spike S1 drive clade success in SARS-CoV-2. bioRxiv. 2021.

6. Goel RR, Painter MM, Apostolidis SA, Mathew D, Meng W, Rosenfeld AM, et al. mRNA Vaccination Induces Durable Immune Memory to SARS-CoV-2 with Continued Evolution to Variants of Concern. bioRxiv. 2021.

7. Scobie HM, Johnson AG, Suthar AB, Severson R, Alden NB, Balter S, et al. Monitoring Incidence of COVID-19 Cases, Hospitalizations, and Deaths, by Vaccination Status - 13 U.S. Jurisdictions, April 4-July 17, 2021. MMWR Morb Mortal Wkly Rep. 2021;70(37):1284–90.

8. Kislaya I, Rodrigues EF, Borges V, Gomes JP, Sousa C, Almeida JP, et al. Delta variant and mRNA Covid-19 vaccines effectiveness: higher odss of vaccine breakthroughs. medRxiv. 2021.

9. Saad-Roy CM, Morris SE, Metcalf CJE, Mina MJ, Baker RE, Farrar J, et al. Epidemiological and evolutionary considerations of SARS-CoV-2 vaccine dosing regimes. Science. 2021;372(6540):363–70.

10. Fauver JR, Petrone ME, Hodcroft EB, Shioda K, Ehrlich HY, Watts AG, et al. Coast-to-Coast Spread of SARS-CoV-2 during the Early Epidemic in the United States. Cell. 2020;181(5):990–6 e5.

11. Oude Munnink BB, Nieuwenhuijse DF, Stein M, O’Toole A, Haverkate M, Mollers M, et al. Rapid SARS-CoV-2 whole-genome sequencing and analysis for informed public health decision-making in the Netherlands. Nat Med. 2020.

12. Gonzalez-Reiche AS, Hernandez MM, Sullivan MJ, Ciferri B, Alshammary H, Obla A, et al. Introductions and early spread of SARS-CoV-2 in the New York City area. Science. 2020;369(6501):297–301.

13. Moreno GK, Braun KM, Riemersma KK, Martin MA, Halfmann PJ, Crooks CM, et al. Distinct patterns of SARS-CoV-2 transmission in two nearby communities in Wisconsin, USA. medRxiv. 2020.

14. Rockett RJ, Arnott A, Lam C, Sadsad R, Timms V, Gray KA, et al. Revealing COVID-19 transmission in Australia by SARS-CoV-2 genome sequencing and agent-based modeling. Nat Med. 2020.

15. Taboada B, Vazquez-Perez JA, Munoz Medina JE, Ramos Cervantes P, Escalera-Zamudio M, Boukadida C, et al. Genomic Analysis of Early SARS-CoV-2 Variants Introduced in Mexico. J Virol. 2020.

16. Candido DS, Claro IM, de Jesus JG, Souza WM, Moreira FRR, Dellicour S, et al. Evolution and epidemic spread of SARS-CoV-2 in Brazil. Science. 2020.

17. Jangra S, Ye C, Rathnasinghe R, Stadlbauer D, Krammer F, Simon V, et al. The E484K mutation in the SARS-CoV-2 spike protein reduces but does not abolish neutralizing activity of human convalescent and post-vaccination sera. medRxiv. 2021.

18. Greaney AJ, Loes AN, Crawford KHD, Starr TN, Malone KD, Chu HY, et al. Comprehensive mapping of mutations in the SARS-CoV-2 receptor-binding domain that affect recognition by polyclonal human plasma antibodies. Cell Host Microbe. 2021;29(3):463–76 e6.

19. Rockett RJ, Arnott A, Lam C, Sadsad R, Timms V, Gray KA, et al. Revealing COVID-19 transmission in Australia by SARS-CoV-2 genome sequencing and agent-based modeling. Nat Med. 2020;26(9):1398–404.

20. Grifoni A, Weiskopf D, Ramirez SI, Mateus J, Dan JM, Moderbacher CR, et al. Targets of T Cell Responses to SARS-CoV-2 Coronavirus in Humans with COVID-19 Disease and Unexposed Individuals. Cell. 2020;181(7):1489–501 e15.

21. Agerer B, Koblischke M, Gudipati V, Montano-Gutierrez LF, Smyth M, Popa A, et al. SARS-CoV-2 mutations in MHC-I-restricted epitopes evade CD8(+) T cell responses. Sci Immunol. 2021;6(57).

22. Skidmore PT, Kaelin EA, Holland LA, Maqsood R, Wu LI, Mellor NJ, et al. Genomic Sequencing of SARS-CoV-2 E484K Variant B.1.243.1, Arizona, USA. Emerg Infect Dis. 2021;27(10):2718–20.

23. Moustafa AM, Bianco C, Denu L, Ahmed A, Coffin SE, Neide B, et al. Comparative Analysis of Emerging B.1.1.7+E484K SARS-CoV-2 Isolates. Open Forum Infect Dis. 2021;8(7):ofab300.

24. Barton MI, MacGowan SA, Kutuzov MA, Dushek O, Barton GJ, van der Merwe PA. Effects of common mutations in the SARS-CoV-2 Spike RBD and its ligand, the human ACE2 receptor on binding affinity and kinetics. Elife. 2021;10.

25. Li Q, Nie J, Wu J, Zhang L, Ding R, Wang H, et al. SARS-CoV-2 501Y.V2 variants lack higher infectivity but do have immune escape. Cell. 2021;184(9):2362–71 e9.

26. Li X, Giorgi EE, Marichann MH, Foley B, Xiao C, Kong XP, et al. Emergence of SARS-CoV-2 through Recombination and Strong Purifying Selection. bioRxiv. 2020.

27. Zhang L, Jackson CB, Mou H, Ojha A, Rangarajan ES, Izard T, et al. The D614G mutation in the SARS-CoV-2 spike protein reduces S1 shedding and increases infectivity. bioRxiv. 2020.

28. Yurkovetskiy L, Pascal KE, Tompkins-Tinch C, Nyalile T, Wang Y, Baum A, et al. SARS-CoV-2 Spike protein variant D614G increases infectivity and retains sensitivity to antibodies that target the receptor binding domain. bioRxiv. 2020.

29. Daniloski Z, Guo X, Sanjana NE. The D614G mutation in SARS-CoV-2 Spike increases transduction of multiple human cell types. bioRxiv. 2020.

30. Kraemer MUG, Hill V, Ruis C, Dellicour S, Bajaj S, McCrone JT, et al. Spatiotemporal invasion dynamics of SARS-CoV-2 lineage B.1.1.7 emergence. Science. 2021;373(6557):889–95.

31. Volz E, Mishra S, Chand M, Barrett JC, Johnson R, Geidelberg L, et al. Assessing transmissibility of SARS-CoV-2 lineage B.1.1.7 in England. Nature. 2021;593(7858):266–9.

32. Annavajhala MK, Mohri H, Wang P, Nair M, Zucker JE, Sheng Z, et al. Emergence and expansion of SARS-CoV-2 B.1.526 after identification in New York. Nature. 2021.

33. Everett J, Hokama P, Roche AM, Reddy S, Hwang Y, Kessler L, et al. SARS-CoV-2 Genomic Variation in Space and Time in Hospitalized Patients in Philadelphia. mBio. 2021;12(1).

34. Sherrill-Mix S, Hwang Y, Roche AM, Glascock A, Weiss SR, Li Y, et al. Detection of SARS-CoV-2 RNA using RT-LAMP and molecular beacons. Genome Biol. 2021;22(1):169.

35. Hilaire BGS, Durand NC, Mitra N, Pulido SG, Mahajan R, Blackburn A, et al. A rapid, low cost, and highly sensitive SARS-CoV-2 diagnostic based on whole genome sequencing. bioRxiv. 2020.

36. Zahradnik J, Marciano S, Shemesh M, Zoler E, Harari D, Chiaravalli J, et al. SARS-CoV-2 variant prediction and antiviral drug design are enabled by RBD in vitro evolution. Nat Microbiol. 2021;6(9):1188–98.

37. Zhu X, Mannar D, Srivastava SS, Berezuk AM, Demers JP, Saville JW, et al. Cryo-electron microscopy structures of the N501Y SARS-CoV-2 spike protein in complex with ACE2 and 2 potent neutralizing antibodies. PLoS Biol. 2021;19(4):e3001237.

38. Farinholt T, Doddapaneni H, Qin X, Menon V, Meng Q, Metcalf G, et al. Transmission event of SARS-CoV-2 Delta variant reveals multiple vaccine breakthrough infections. medRxiv. 2021.

39. Kustin T, Harel N, Finkel U, Perchik S, Harari S, Tahor M, et al. Evidence for increased breakthrough rates of SARS-CoV-2 variants of concern in BNT162b2-mRNA-vaccinated individuals. Nat Med. 2021;27(8):1379–84.

40. Mlcochova P, Kemp S, Dhar MS, Papa G, Meng B, Ferreira I, et al. SARS-CoV-2 B.1.617.2 Delta variant replication and immune evasion. Nature. 2021.

41. Lopez Bernal J, Andrews N, Gower C, Gallagher E, Simmons R, Thelwall S, et al. Effectiveness of Covid-19 Vaccines against the B.1.617.2 (Delta) Variant. N Engl J Med. 2021;385(7):585–94.

42. McEwen AE, Cohen S, Bryson-Cahn C, Liu C, Pergam SA, Lynch J, et al. Variants of concern are overrepresented among post-vaccination breakthrough infections of SARS-CoV-2 in Washington State. Clin Infect Dis. 2021.

43. Martin DP, Weaver S, Tegally H, San JE, Shank SD, Wilkinson E, et al. The emergence and ongoing convergent evolution of the SARS-CoV-2 N501Y lineages. Cell. 2021;184:5189–200.

44. McCallum M, De Marco A, Lempp FA, Tortorici MA, Pinto D, Walls AC, et al. N-terminal domain antigenic mapping reveals a site of vulnerability for SARS-CoV-2. Cell. 2021;184(9):2332–47 e16.

45. Syed AM, Taha TY, Khalid MM, Tabata T, Chen IP, Sreekumar B, et al. Rapid assessment of SARS-CoV-2 evolved variants using virus-like particles. bioRxiv. 2021.

46. St. Hilaire BG, Durand NC, Mitra N, Pulido SG, Mahajan R, Blackburn A, et al. A rapid, low cost, and highly sensitive SARS-CoV-2 diagnostic based on whole genome sequencing. bioRxiv. 2020.

47. Li H, Durbin R. Fast and accurate short read alignment with Burrows-Wheeler transform. Bioinformatics. 2009;25(14):1754–60.

48. Li H, Handsaker B, Wysoker A, Fennell T, Ruan J, Homer N, et al. The Sequence Alignment/Map format and SAMtools. Bioinformatics. 2009;25(16):2078–9.

49. Li H. A statistical framework for SNP calling, mutation discovery, association mapping and population genetical parameter estimation from sequencing data. Bioinformatics. 2011;27(21):2987–93.

50. Carpenter B, Gelman A, Hoffman MD, Lee D, Goodrich B, Betancourt M, et al. STAN: A Probabilisitic Programming Language. J Stat Software. 2017;76:1–32.

