## Supplemental Table 2 for "SARS-CoV-2 variants associated with vaccine breakthrough in the Delaware Valley through summer 2021"

**Supplementary Table 3:** Nomenclature of Variants Being Monitored and Variants of Concern

**Pango Lineage	WHO Designation	VBM/VOC	Location of First Identification**

B.1.1.7	Alpha	VBM	United Kingdom

Q.1	Alpha	VBM

Q.4	Alpha	VBM

Q.6	Alpha	VBM

B.1.351	Beta	VBM	South Africa

P.1	Gamma	VBM	Brazil/Japan

P.1.10	Gamma	VBM

P.1.2	Gamma	VBM

B.1.427	Epsilon	VBM	California, USA

B.1.429	Epsilon	VBM	California, USA

B.1.525	Eta	VBM	United Kingdom/Nigeria

B.1.526	Iota	VBM	New York, USA

B.1.617.1	Kappa	VBM	India

B.1.621	Mu	VBM	Columbia

B.1.621.1	Mu	VBM

P.2	Zeta	VBM	Brazil

B.1.617.2	Delta	VOC	India

AY.1	Delta	VOC

AY.10	Delta	VOC

AY.12	Delta	VOC

AY.13	Delta	VOC

AY.14	Delta	VOC

AY.15	Delta	VOC

AY.19	Delta	VOC

AY.2	Delta	VOC

AY.20	Delta	VOC

AY.21	Delta	VOC

AY.24	Delta	VOC

AY.25	Delta	VOC

AY.3	Delta	VOC

AY.3.1	Delta	VOC

AY.4	Delta	VOC

AY.5	Delta	VOC

AY.6	Delta	VOC
