## Supplemental Table 8 for "SARS-CoV-2 variants associated with vaccine breakthrough in the Delaware Valley through summer 2021"

**Supplementary Table 8:** Key Reagents

**Reagent or Resource	Source	Identifier**

QIAmp 96 Viral RNA Kit	Qiagen, Hilden, Germany	5262

SupperScript III RT	Thermo Fisher Scientific, Waltham, USA	56575

SS III First Strand 5x Buffer	Thermo Fisher Scientific, Waltham, USA	Y02321

Random Hexamers	Thermo Fisher Scientific, Waltham, USA	51709

Dithiothreitol	Thermo Fisher Scientific, Waltham, USA	Y00122

Molecular Grade Water	Thermo Fisher Scientific, Waltham, USA	Y01138

Deoxynucleotide Mix	New England Biolabs, Ipswich, USA	N0447S

ARTIC Primer Pool 1	Integrated DNA Technologies, Coralville, USA	100006786

ARTIC Primer Pool 2	Integrated DNA Technologies, Coralville, USA	100006787

Q5 Hot Start Polymerase	New England Biolabs, Ipswich, USA	M0493L

Q5 5x Reaction Buffer	New England Biolabs, Ipswich, USA	B90275

AMPure XP	Beckman Coulter, Brea, USA	A63882

Qubit™ 1X dsDNA Kit	Invitrogen Corp., Waltham, USA	Q33230

Quant-iT PicoGreen Kit	Invitrogen Corp., Waltham, USA	P7589

IDT for Illumina DNA/RNA	Illumina Inc., San Diego, USA	20027213

UD Indexes A-D	20027214

20027215

20027216

Nextera XT DNA Library	Illumina Inc., San Diego, USA	FC-131-1096

Preparation Kit

NextSeq 500/550 Mid Output	Illumina Inc., San Diego, USA	20024904

Kit v2.5 (150 Cycles)
